# Validation and reproducibility of mapping positron emission tomography uptake in the aortic wall and thrombus

**DOI:** 10.1101/2024.07.30.24311215

**Authors:** Mireia Bragulat-Arévalo, Marta Ferrer-Cornet, Lydia Dux-Santoy, Ruper Oliveró-Soldevila, Marvin Garcia-Reyes, Gisela Teixido-Tura, Juan Garrido-Oliver, Laura Galian-Gay, Pere Lopez-Gutierrez, Alberto Morales-Galán, Alba Catalá-Santarrufina, Jose Ramon García Garzón, Noemi Martinez Esquerda, Javier Solsona, Ignacio Ferreira-González, Sergi Bellmunt-Montoya, Jose Rodriguez-Palomares, Andrea Guala

**Author notes:** Equally contributed (Mireia Bragulat-Arévalo, Marta Ferrer-Cornet). **Corresponding authors:** Lydia Dux-Santoy and Jose Rodriguez-Palomares, Vall d’Hebron Institut de Recerca (VHIR), Barcelona, Spain. Mails.

## Abstract

**INTRODUCTION:** Aortic aneurysms are prevalent diseases of the aorta, with limited knowledge about their aetiology, progression, and risk, and no effective pharmacological treatment. 18F-fluorodesoxyglucose Positron Emission Tomography (18F-FDG PET) provides molecular level information of glucose activity, serving as potential analogue of vascular inflammation, which may be relevant to improve knowledge and clinical management of aortic aneurysms. Nonetheless, current clinical assessment of 18F-FDG uptake presents several limitations.

**PURPOSE:** The aim of this study was to develop, test and validate an innovative aortic wall uptake mapping.

**METHODS:** PET/magnetic resonance (MR) data from 72 patients were acquired. The aortic lumen was segmented, and 9 anatomical landmarks were identified to create a standardised aortic lumen discretization including up to 80 patches. Black-blood MR images were used to guide thrombus segmentation if present, which was merged with lumen segmentation to generate a thoraco-abdominal aortic wall mask. This mask was then expanded by 1 to 5 mm in the inward and outward directions, resulting in an aortic wall volume of interest. Median and percentile 95th standard uptake value (SUV) was calculated on each aortic patch. A second observer performed the same analysis on 23 randomly selected patients for inter-observer reproducibility assessment. Validation was performed by comparisons with regional SUV measured by a nuclear medicine expert.

**RESULTS:** The technique was highly feasible, permitting the analysis of all 72 patients and in the 99.6% of aortic wall patches. The image analysis workflow was highly reproducible, resulting in Dice scores of 0.90 [0.57, 0.91] and 0.85 [0.78, 0.88] for aortic and thrombus segmentations, respectively, and in excellent co-localization for anatomical references (5.74 [3.62, 8.73] mm).

The inter-observer reproducibility of aortic wall SUV mapping was excellent (ICC between 0.924 and 0.945), with limited differences with respect to aortic wall thickness and similar performances for SUV quantification in the thrombus. The validation of regional SUV values showed good agreement, with limited impact of aortic wall thickness values. A balance between reproducibility and accuracy was obtained with a volume of interest of 6 mm thickness.

**CONCLUSIONS:** An image analysis implementation based on multi-modality PET/MR data provides reproducible and accurate quantitative aortic wall 18F-FDG uptake maps. This approach may enable exploring local factors related to an inflammatory vascular state, with possible repercussions in clinical practice.

## Introduction

Aortic aneurysms are a pathological dilation of the aorta and one of the most frequent disease of the aorta, with an incidence of 3.5 to 6.5 / 1000 person [1]. Aortic aneurysms are mostly asymptomatic, which complicates their diagnosis, the control of patients evolution, and increases the risk of two life threatening cardiovascular events, aortic rupture and dissection [2]. As no drug has been proven effective to halt or reduce aneurysms growth, the only viable treatment is surgical or endovascular intervention, which have inherent risks [1]. Diagnosis, risk-stratification and the decision for intervention is based on maximum aortic diameter [3]. Although it is a well-established method, it has limited capacity for the prediction of rupture [4][5] and suffers from observer variability [6]. Thus, new biomarkers of aortic risk as well as new treatments are urgently needed [7].

The aetiology of the vast majority of aortic aneurysms is unknown, which limits the identification and development of effective pharmacological treatments [7], [8]. Certain studies have reported inflammatory cells influx in the aortic wall as an early event in aneurysm formation [9], [10], [11]. 18F-fluorodesoxyglucose Positron Emission Tomography (18F-FDG PET) could be a useful imaging technique to assess aortic aneurysm inflammation, as it provides molecular level information of glucose activity, commonly used for the detection of inflammation [12][13]. Importantly, a strong correlation between 18F-FDG activity and atherosclerotic disease was reported [14], but it did not clearly translate to aneurysm growth and risk prediction [9] [10], while a possible role of thrombus-driven hypoxia was established [15]. The heterogeneity of results has been attributed to the lack of cell-specificity absorption of the 18F-FDG in the macrophages involved [16] and may be related to methodological differences in PET analysis [9] and limited capacity to measure aneurysms growth [17]. Notably, most studies suggested further analysis on larger cohorts [10], [11].

In current clinical practice and research, the analysis of PET images lacks solid standardization, and is mostly performed visually (qualitative). When done semi-quantitatively, nuclear medicine physicians extract the maximum standard uptake value (SUV) by manually drawing a region of interest where the highest activity is visualized [18], which improves diagnostic performance compared to visual analysis [19] but presents limited reproducibility [20]. Moreover, this methodology might be insufficient to quantify sub-clinical inflammation [9] and does not permit to assess spatially resolved activity, which may be relevant in aortic disease. A recent study proposed the mapping of PET uptake over the aortic wall, showing excellent reproducibility, but a formal validation remain to be demonstrated [21].

This study aimed to develop and validate an image analysis technique to extract 18F-FDG uptake maps over the thoraco-abdominal aortic wall from PET/magnetic resonance (MR) equipment, and to assess its inter-observer reproducibility.

## Methodology

### Study cohort

As part of 2 ongoing projects, PET/MR scans from 72 adult patients (age > 18 years) with aortic aneurysms of non-genetic aetiology, without previous aortic surgery or history of acute aortic syndromes, and free from severe aortic valve disease and contraindications for MR or PET were prospectively acquired. Among them, 43 patients had a thoracic descending or abdominal aortic aneurysm with a maximum diameter ≥ 40 and <55 mm while the remaining 29 patients had a bicuspid aortic valve and a maximum diameter of the ascending aorta < 50 mm. The 43 patients with disease involving the thoracic descending or abdominal aorta underwent PET/MR imaging of the whole thoracic-abdominal aorta, while the 29 with ascending aorta diseases underwent imaging of only the thoracic region. This study was approved by the Vall d’Hebron University Hospital Ethics committee, and patients gave their written informed consent.

### Image Acquisition

All acquisitions were performed in a hybrid whole-body PET/MR scanner (SIGNA™ PET/MR, General Electric HealthCare). MR acquisitions were performed at 3T and included (i) a 4D flow MR sequence covering the thoracic or thoracic-abdominal aorta field of view (FOV) (homogeneous pixel size of 1.64 [1.41, 1.72] mm), (ii) a non-ECG gated, contrast-enhanced magnetic resonance angiography (MRA) with administration of gadolinium chelated contrast (homogeneous pixel size of 0.82 [0.82, 0.86] mm) and (iii) a blood-suppressed single-shot fast-spin echo (SSFSE) sequence [22]. Data from the 4D flow sequence was used to reconstruct a phase-contrast enhanced MRA (PC-MRA) following [23].

For PET imaging, all patients were administered 4 MBq/kg of 18F-FDG intravenously 90 minutes before PET acquisition. Attenuation correction was based on fast spoiled gradient echo MR sequences which generates 4 contrast maps in fast acquisition of only water, fat, in-phase and out-phase (Dixon sequences). Image reconstruction was performed automatically using time of flight algorithm (Q.Clear, General Electric HealthCare), which reduces noise (homogeneous pixel size of 3.13 [3.13, 3.13] mm). SUV was computed as the decay corrected 18F-FDG concentration divided by the injected dose and patient’s body weight [24].

### Image Analysis

An overview of the image analysis is included in Figure 1. The thoracic aorta was segmented semi-automatically on PC-MRA images while the abdominal aorta was segmented on MRA data using 3D Slicer [25]. Thrombus was segmented on MRA images guided by SSFSE sequence. For the patients with thoracic-abdominal FOV and thrombus, all segmentations were merged generating a unique 3D model, a task that was not required in patients with thoracic FOV. Then, the aortic centerline was generated, and 9 anatomical landmarks (aortic annulus, sinotubular junction, brachiocephalic artery, left subclavian artery, diaphragmatic level, pulmonary bifurcation level, superior mesenteric artery, renal arteries, aortoiliac bifurcation) were identified (Fig. 1.2). The precision of the colocalization between MR segmentations and PET images was confirmed visually, and manual correction of misalignments was performed by rigid transformation if needed (Fig. 1.3). To test the inter-observer reproducibility, the whole annotation procedure was repeated by a second observer, blinded to the other observer annotations, in 23 randomly identified patients.

**Figure 1.**
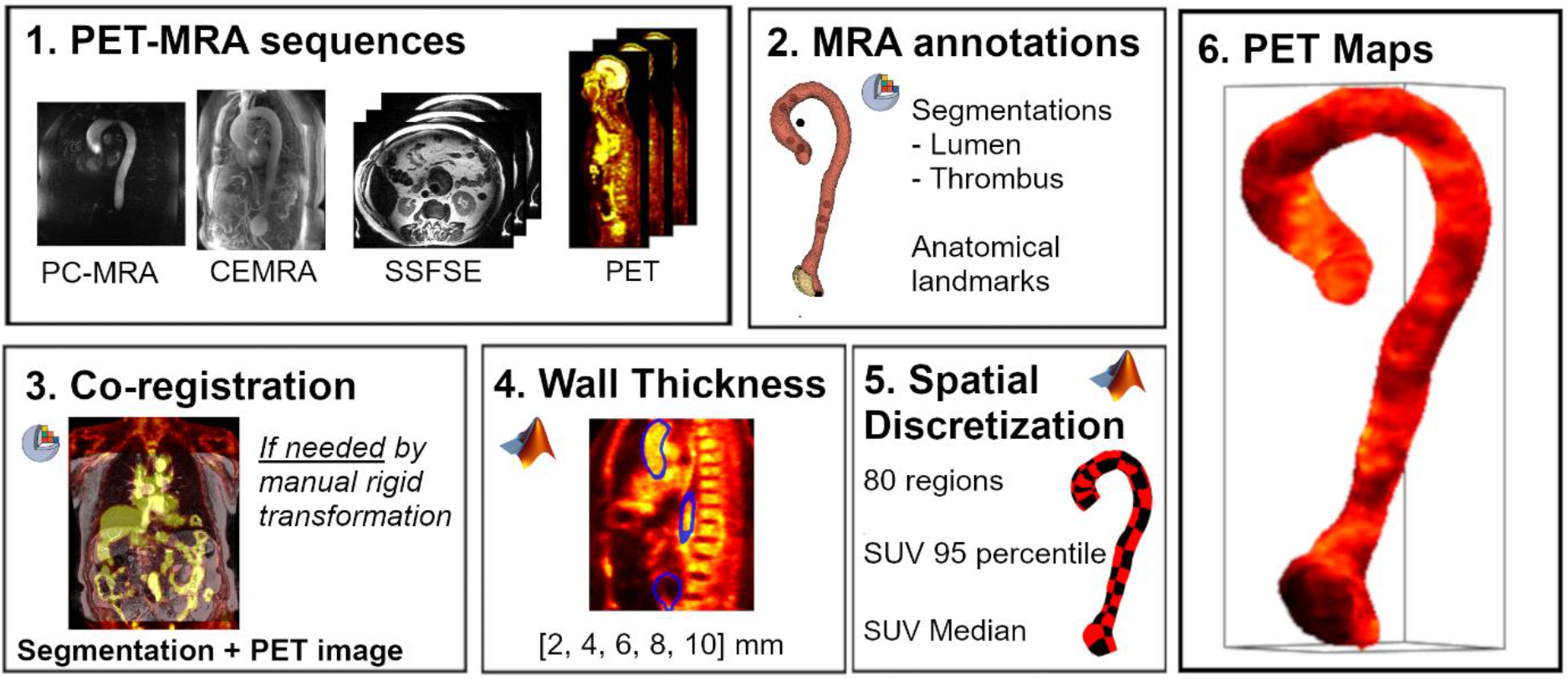
Overview of the image analysis

The subsequent image analysis was performed in Matlab (The Mathworks, Natick, MA, USA). The surface of the 3D segmentation of the aorta underwent an inward and outward expansion to create an aorta wall volume, which was voxelized with a uniform spatial resolution of 1 mm. To test the impact of wall thickness in subsequent extraction, five different thicknesses (from 1 to 5 mm on each side) were tested (Fig. 1.4). SUV of PET voxels were interpolated to match this resolution. Using a hot color-coded scale (black the lowest and white the highest), aortic mapping of the SUV was reconstructed (Fig. 1.6). Performing a visual analysis of each patient’s map, spill-over from the myocardium and other artifacts were identified: if minor and very local, segmentations were corrected before further analysis, while if the artifact was substantial those pixels affected were manually removed from the analysis. The aortic wall volume was then discretized into 52 (thoracic FOV) or 80 (thoracic-abdominal FOV) wall patches. To this aim, using the 9 anatomical points and the centerline, the ascending aorta, aortic arch, and thoracic and abdominal descending aorta were discretized into 4, 2, 7 and 7 longitudinal regions, respectively. Then, each longitudinal region was divided into 4 circumferential patches (inner, right, outer, left), generating the aortic patches (Fig. 1.5). For each region, the median and 95^th^ percentile SUV were calculated. Additionally, the median and 95^th^ percentile SUV for the thrombus volume were computed.

### Validation

Clinical evaluation of PET images was performed by a nuclear medicine physician (JRGG) with extensive experience in PET imaging using Carestream PACS Client Suite (Carestream, Rochester, NY). The physician measured maximum SUV uptake in the ascending, thoracic-descending, and abdominal aorta, and in the aneurysmal region following current practice. For the validation test, and to match the clinical differentiation of the aorta, the maximum SUV obtained by the mapping technique in the same regions were computed.

### Statistical Analysis

Inter-observer reproducibility as well as validation of aortic wall SUV maps was tested for the five wall thicknesses investigated. Inter-observer reproducibility for the segmentations was evaluated in terms of the 95% Hausdorff Distance (HD) and Dice Score Coefficient (DSC), while absolute distance was used for the anatomical landmarks. To assess the SUV mapping reproducibility, the median and 95th percentile SUV for each aortic patch as well as those of the thrombus obtained by the two observers were compared via intra-class correlation coefficient (ICC, by two-way random-effects model, single rater and absolute agreement) [26]. On the other hand, to validate the methodology, the maximum SUV of the ascending, descending and abdominal aorta, and aneurysmal region from both observers on each thickness was compared with the SUV reported by the expert nuclear medicine physician by linear regression and Bland-Altman analysis.

Continuous variables with normal distribution were presented as mean ± standard deviation, while for non-normally distributed variables median and inter-quartile [IQR] range were used. Categorical variables were presented as number (percentage). All statistical analyses were performed using R Studio (R: A Language and Environment for Statistical Computing, R Core Team, Vienna, Austria).

## Results

Seventy-two patients were included in the analysis. Demographic and clinical variables are summarized in Table 1. A total of 5032 aortic patches were extracted (3744 in the thoracic, 1288 in the abdominal region). Two PET images were affected by artifacts while PET images of 4 patients presented signal spill-over from the myocardium: these artifacts required manual correction of a total of 20 (0.3 %) patches. Thus, the feasibility of the patch-wise PET analysis was 99.6%.

**Table 1.** Patient characteristics.

| Characteristic | N = 72 |
| --- | --- |
| Age (years) | 67 [56, 74] |
| Male | 56 (77.8 %) |
| Body surface area (m<sup>2</sup>) | 1.91 ± 0.22 |
| Aortic Valve |  |
| *Bicuspid aortic valve* | 29 (40.3) |
| *Tricuspid aortic valve* | 43 (59.7 %) |
| Aneurysm Location |  |
| *Thoracic Ascending* | 32 (44.4 %) |
| *Thoracic Descending* | 4 (5.6 %) |
| *Abdominal* | 34 (47.2 %) |
| Thrombus | 28 (39.4 %) |

### Inter-observer reproducibility

Excellent reproducibility was obtained for the aortic segmentation (HD 3.41 [2.59, 4.35] mm, DSC 0.90 [0.57, 0.91]) and thrombus segmentation (HD 5.16 [3.5, 8.8] mm, DSC 0.85 [0.78, 0.88]). Similarly, good reproducibility was obtained for the localization of the anatomical landmarks (5.74 [3.62, 8.73] mm). The inter-observer reproducibility of aortic SUV mapping was excellent, with ICC in between 0.924 and 0.945 (Figure 2 and 3, Table 2). Comparing inter-observer reproducibility on aortic wall volume of different thicknesses, little difference was found, with a tendency of higher reproducibility at the largest thickness value (SUV median ICC increases from 0.924 at 2 mm to 0.940 at 10 mm). Similarly, good inter-observer reproducibility was obtained for the assessment of median (ICC = 0.890 [0.698, 0.963]) and 95^th^ percentile (ICC = 0.735 [0.363, 0.906]) SUV in the thrombus (Figure 4).

**Table 2:** Inter-observer reproducibility and validation of aortic wall SUV mapping at different wall thicknesses. Bold represents the best values for each parameter among all thicknesses.

| Thickness |  | 2 [mm] | 4 [mm] | 6 [mm] | 8 [mm] | 10 [mm] |
| --- | --- | --- | --- | --- | --- | --- |
| **Interobserver Reproducibility** |  |  |  |  |  |  |
| **R** | Median | 0.924 | 0.929 | 0.935 | 0.936 | **0.940** |
|  | 95th percentile | 0.924 | 0.930 | 0.937 | 0.936 | **0.946** |
| **Intercept** | Median | 0.075 | 0.072 | 0.066 | 0.064 | **0.063** |
|  | 95th percentile | 0.168 | 0.155 | 0.147 | 0.158 | **0.137** |
| **Slope** | Median | 0.951 | 0.953 | 0.957 | 0.959 | **0.960** |
|  | 95th percentile | 0.916 | 0.922 | 0.926 | 0.922 | **0.933** |
| **ICC** | Median | 0.924<br>[0.916, 0.931] | 0.929<br>[0.922, 0.935] | 0.934<br>[0.928, 0.940] | 0.936<br>[0.930, 0.942] | **0.940**<br>**[0.934, 0.945]** |
|  | 95th percentile | 0.924<br>[0.917, 0.931] | 0.930<br>[0.923, 0.936] | 0.936<br>[0.930, 0.942] | 0.936<br>[0.930, 0.942] | **0.945**<br>**[0.940, 0.950]** |
| **Validation with manual assessment** |  |  |  |  |  |  |
| **R** | Maximum | 0.696 | **0.699** | 0.688 | 0.661 | 0.615 |
| **Intercept** | Maximum | **0.5** | 0.519 | 0.555 | 0.66 | 0.761 |
| **Slope** | Maximum | 0.939 | 0.952 | **0.979** | 0.964 | 0.963 |
| **ICC** | Maximum | **0.62**<br>**[0.44, 0.74]** | 0.607<br>[0.386, 0.74] | 0.56<br>[0.255, 0.728] | 0.516<br>[0.188, 0.701] | 0.45<br>[0.124, 0.649] |

**Figure 2.**
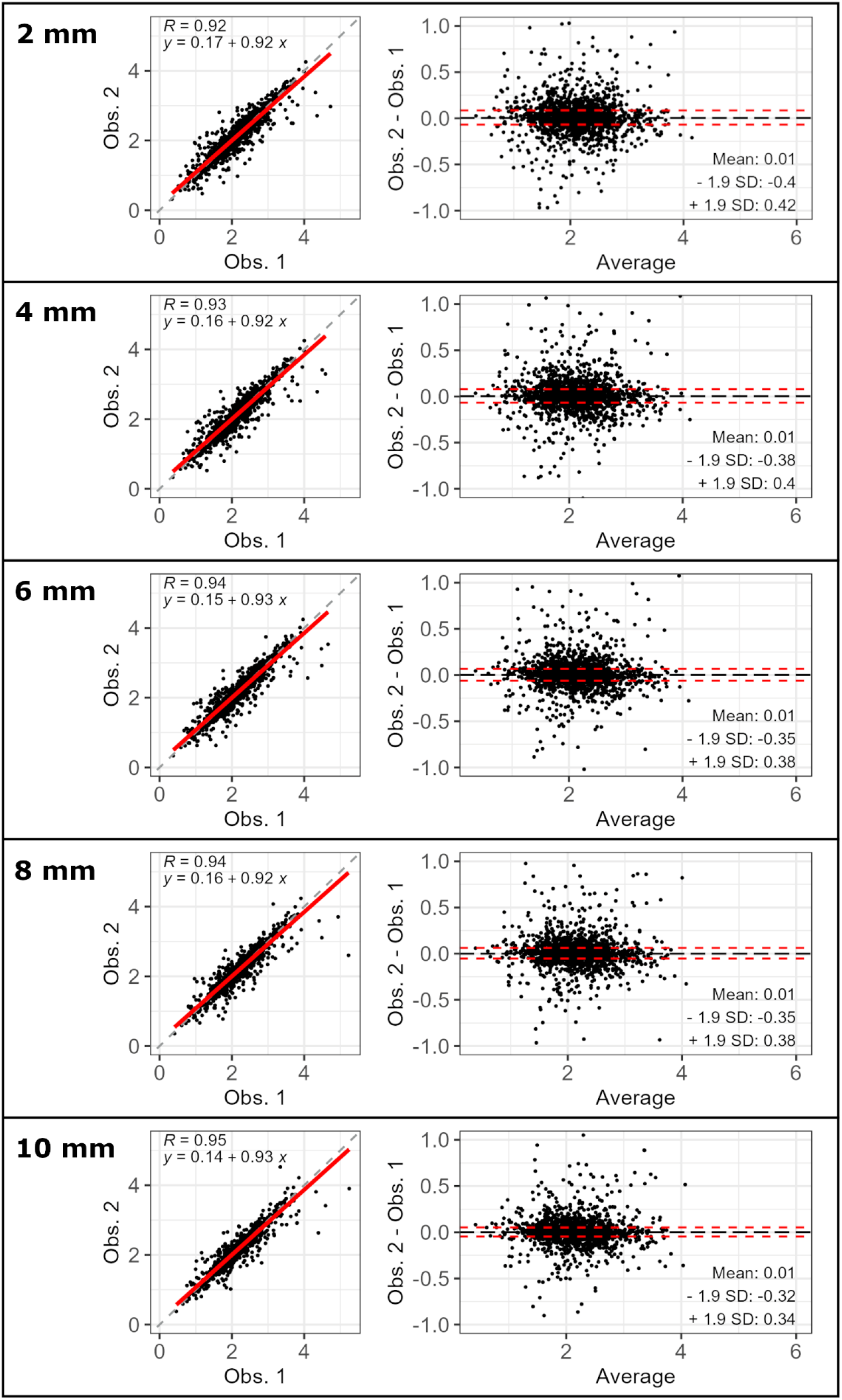
Scatter (left) and Bland-Altmann (right) plots for the inter-observer variability of 95^th^ percentile SUV maps according to aortic wall thickness (2 to 10 mm, top-down order)

**Figure 3.**
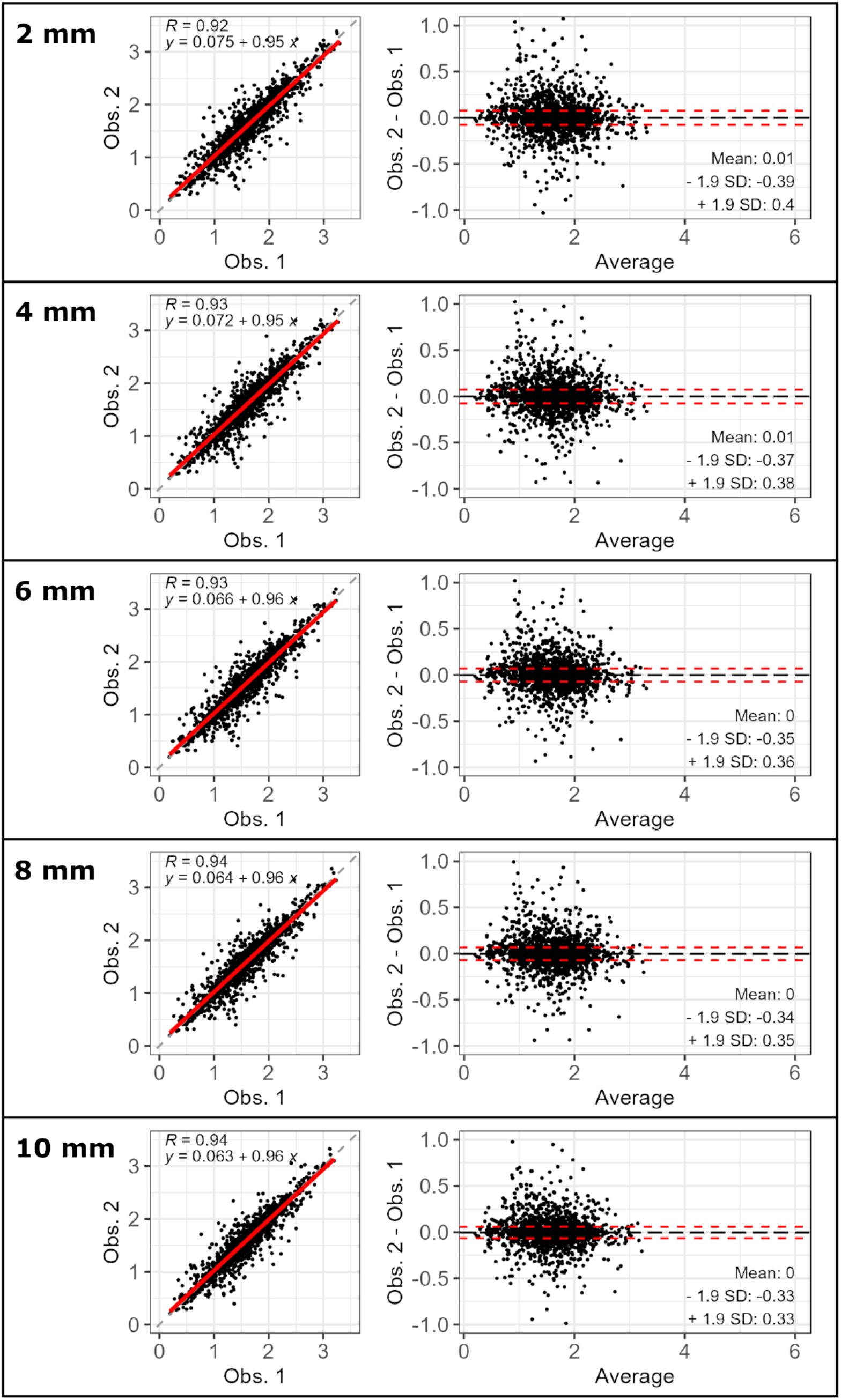
Scatter (left) and Bland-Altmann (right) plots for the inter-observer variability of median SUV maps according to aortic wall thickness (2 to 10 mm, top-down order)

**Figure 4.**
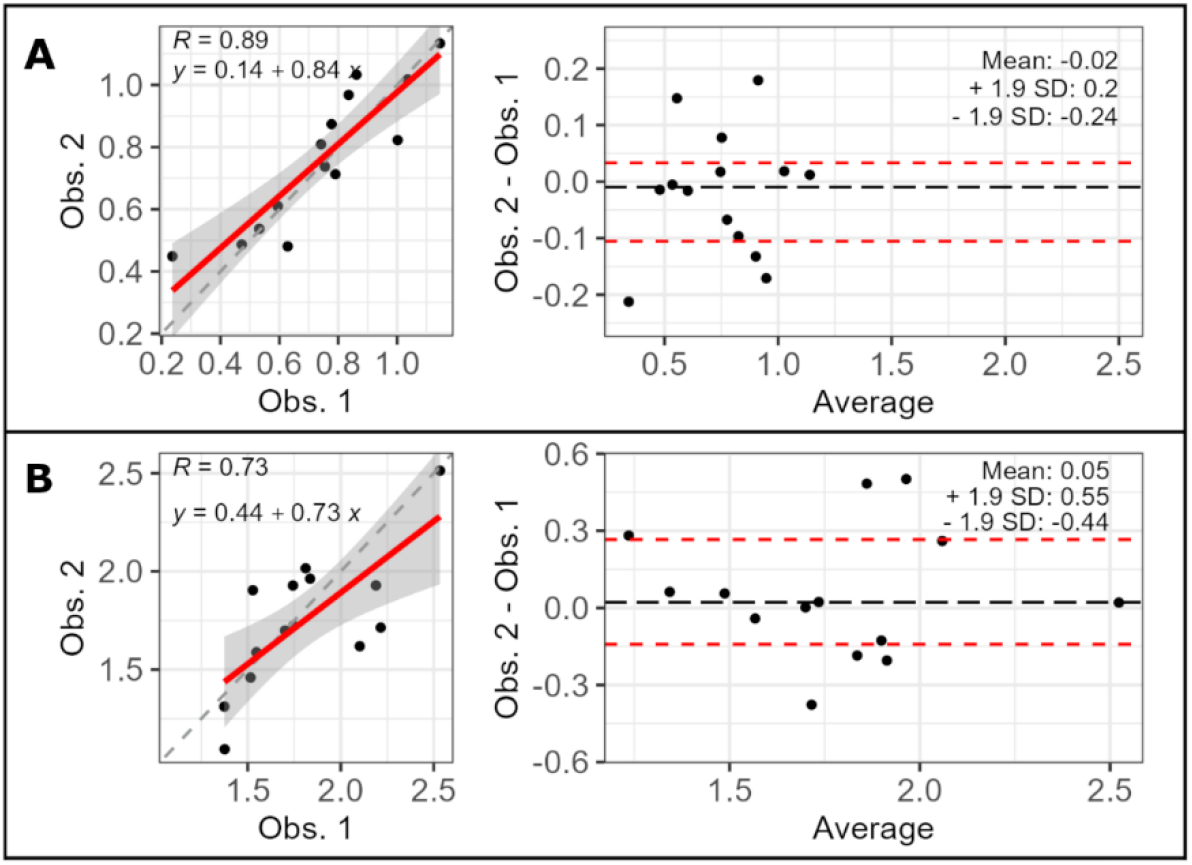
Inter-observer reproducibility of median (A) and 95^th^ percentile (B) SUV uptake in the thrombus.

### Validation

A validation of regional maximum SUV obtained by the mapping technique was performed by comparison against 224 data points manually assessed by an expert. Results showed good agreement with reference standard (linear correlation coefficient R between 0.615 at 10 mm and 0.686 at 2 mm), with little impact of different aortic wall thickness values (Figure 5 and Table 2). The limited differences with respect to aortic wall thickness slightly favoured the thinnest aortic wall volumes.

**Figure 5.**
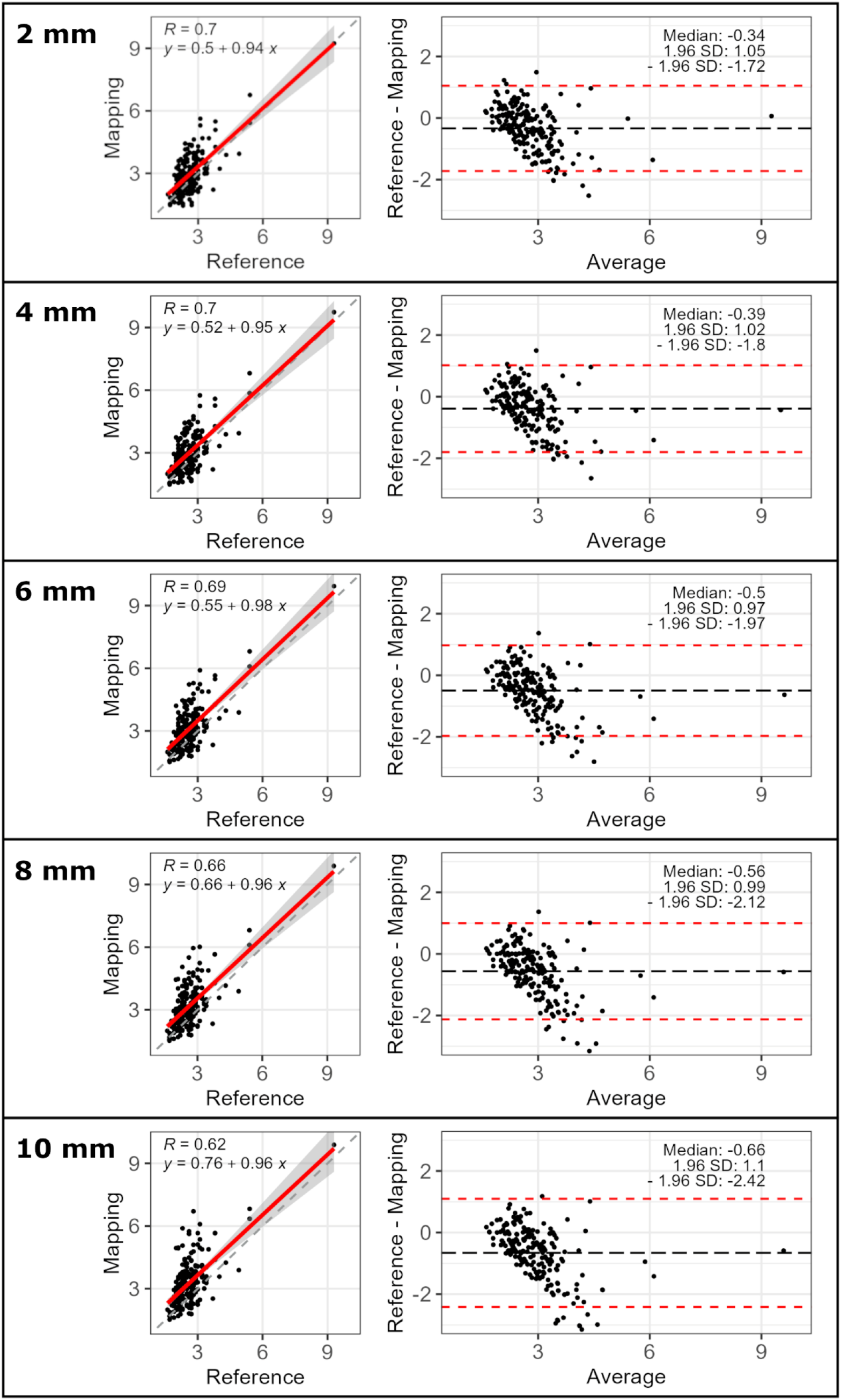
Scatter (left) and Bland-Altmann (right) plots for the validation of maximum SUV values according to aortic wall thickness (2 to 10 mm, top-down order)

## Discussion

In the present study a semi-automatically method for 18F-FDG PET uptake mapping on the aortic wall is presented. Different thicknesses for the aortic wall have been tested to identify possible impact on reproducibility and accuracy. Results showed that the method is highly feasible, reproducible and accurate, and thus is well-suited to assess the eventual role of aortic wall inflammation and its effect in the development and progression of aortic diseases.

PET/MR is a recently introduced hybrid imaging modality allowing for simultaneous acquisition of PET and MR images. These scanners allow for a comprehensive and multimodal analysis of the aorta, permitting simultaneous geometrical, hemodynamic and molecular characterization. Compared to the more established PET/computed tomography scanners, the use of PET/MR results in lower radiation exposure and improved tissue characterization, along with the high flexibility given by the variety of MR sequences available, while maintaining excellent scan-rescan repeatability [27], diagnostic utility [28] and research value [29]. Nonetheless, the full potential of PET/MR in aortic diseases remains to be established. Indeed, current image analysis evaluates the aortic uptake with a unique maximum value, which simplifies the evaluation but is observer dependent and neglects the uptake in other regions as well as its distribution along the aorta. For example, a previous study reported that the number of 18F-FDG hotspots but not their maximum SUV is correlated with aneurysm growth [29], a finding that might be related to the methodology used to quantify SUV or by the importance of SUV distribution. Coping with these limitations, the here-presented implementation permits regional characterization to assess the 3D distribution of inflammatory markers, opening for regional, quantitative analysis and comparisons between segments and co-localised markers of disease severity and progression.

The inter-observer variability in segmentations and anatomic landmarks localization demonstrated minor impact on the calculation of SUV in each patch of the aortic wall. Similarly, very little variation resulted from changing aortic wall thickness. In this respect, inter-observer reproducibility improved slightly at wider wall thicknesses, being 10 mm the best-performing one. At larger wall thickness, patch SUV value is calculated on a thicker aortic wall, which is composed of a larger number of data points: this may mitigate observers discrepancies in the identification of the aortic wall surface. The opposite trend was obtained in the validation analysis, where thinner aortic wall compared better with the reference method. Indeed, increasing aortic wall thickness was associated with an increase in the maximum uptake detected per region, which become slightly more overestimated. We speculate that this may be due to the fact that larger thicknesses may capture more surrounding tissue and organs, and thus the extracted maximum might not be representative of the aortic region of interest. Based on these opposite trends, present results suggest that a wall thickness of 6 mm may be the most favourable balance between accuracy and reproducibility. Notably, the only previous study implementing, but not validating, a similar method used a wall thickness of 6 mm [21].

The reproducibility of results in the thrombus was strong for both inter-observer segmentation and SUV metrics. While not all aneurysms present thrombus, the majority of abdominal aneurysms included in this cohort (39.4 %) presented substantial thrombus. The role of thrombus in modulating the risk of complications in abdominal aortic aneurysms remains a topic of debate among researchers and clinicians [15], [30], [31], [32]. This methodology has the potential to provided more detailed information on the possible interplay between aortic wall metabolism, thrombus presence and aneurysms evolution, which should be the topic of further studies.

## Limitations

The validation of this new technique was limited by the available data obtained as per clinical practice. This resulted in the impossibility to validate the uptake in the thrombus as well as a fully patch-wise validation in the aortic wall. For these analyses, the inter-observer agreement was excellent, leaving little doubt that results are not representative of local PET uptake.

## Conclusions

A technique permitting the 3D mapping of quantitative aortic wall 18-FDG SUV from PET/MR scans was presented, showing excellent reproducibility and accuracy. This technique may improve the understanding of the impact of inflammation on aortic aneurysms etiology and evolution.

## Funding

This work was supported by funding from the Spanish Ministry of Science and Innovation (FORT23/00034, PLEC2021-007664, RTC2019-007280-1), the Instituto de Salud Carlos III (PI19/01480) and “la Caixa” Foundation (LCF/BQ/PR22/11920008).

Acknowledgments

The authors wish to thank Graham Watling for help with the English version of the manuscript.

## Data Availability Statement

All data produced in the present study are available upon reasonable request to the authors.

